# Midlife and old-age cardiovascular risk factors, educational attainment, and cognition at 90-years – population-based study with 48-years of follow-up

**DOI:** 10.1101/2025.03.04.25323082

**Authors:** Anni Varjonen, Toni Saari, Sari Aaltonen, Teemu Palviainen, Mia Urjansson, Paula Iso-Markku, Jaakko Kaprio, Eero Vuoksimaa

## Abstract

We examined the associations of midlife and old-age cardiovascular risk factors, education, and midlife dementia risk scores with cognition at 90+ years, using data from a population-based study with 48 years of follow-up. Participants were 96 individuals aged 90-97 from the older Finnish Twin Cohort study. Cardiovascular risk factors assessed via questionnaires in 1975, 1981, 1990, and 2021-2023 included blood pressure, body mass index, physical activity, and cholesterol, and self-reported educational attainment. The Cardiovascular Risk Factors, Aging, and Dementia (CAIDE) score and an educational-occupational attainment score were used as midlife dementia risk scores. Cognitive assessments included semantic fluency, and immediate and delayed recall from a 10-word list learning task. Regression analyses were conducted with dementia risk factors predicting cognition at 90+ years, adjusting for age, sex, education, follow-up time, and apolipoprotein E genotype (ε4-carrier vs non-carriers). Results showed that higher education and higher educational occupational score were associated with better cognitive performance in all cognitive measures. Those with high midlife blood pressure scored significantly higher in all cognitive tests than those with normal blood pressure. Conversely, those with high old-age blood pressure scored lower in semantic fluency but not in immediate or delayed recall. Other cardiovascular risk factors and the CAIDE score did not show consistent associations with cognition. Education appears to have a long-lasting protective effect in aging, whereas midlife and old-age cardiovascular risk factors showed no consistent associations with cognition in this older population.

## Background

Those who are 90-years or older are the fastest growing population segment in many countries [1], with a substantial proportion having cognitive impairment and high prevalence of dementia, estimated nearly at 40% [2]. Potentially modifiable lifestyle factors are associated with dementia risk differently across the lifespan with seemingly decreasing adverse effects towards older age [3–6]. There is strong evidence for education to serve as a protective factor against dementia [7]. Cardiovascular (CV) risk factors measured in midlife have been associated with increased dementia risk [7,8], but there is evidence that CV risk factors at old age are protective against dementia; for example, higher body mass index (BMI) and developing hypertension at old age have been associated with better old age cognition [3,6,9]. These paradoxical findings likely reflect reverse causation as the risk factors are often measured with short follow-ups, during the preclinical stage of the dementia process which starts even 20 years before the clinical diagnosis [10]. There is a need for studies where CV risk factors have been measured in middle age before the disease process has started, preferably with over 20 years of follow-up to rule out reverse causation [9].

Our aim was to use a population-based sample of twins with up to 48 years of follow-up to investigate if midlife and old age CV risk factors, including BMI, blood pressure (BP), cholesterol, and physical activity (PA) predict cognition at 90 years of age. We also examined education, and two previously validated midlife dementia risk scores, the Cardiovascular Risk Factors, Aging and Dementia (CAIDE) [11], and the educational-occupational score [12] in association with cognition at 90 years old. We hypothesized that those with more midlife CV risk factors, and lower education, have poorer cognition than those with less midlife CV risk factors and higher education. We also expected that the CV risk factors measured at 90 years are not associated or have a reversed association with cognition.

## Materials & methods

### Participants

The participants were twins (irrespective of co-twin’s vital status or participation) who were 90 years old or older, from a population-based older Finnish Twin Cohort (FTC) (established in 1974) [13] and participated in the NONAGINTA –study. The data collection began in June 2020 and is still ongoing. For NONAGINTA enrolment, those who were 90 years old or older from the FTC (born before May 1930) were invited to participate, and invitations were sent when people had reached 90 years of age (those born in 1930-33). Data collection included telephone interviews for cognition and postal questionnaires for lifestyle factors. We also used earlier FTC data that were collected through postal questionnaires with a baseline in 1975 (all twins), and follow-ups in 1981 (all twins) and 1990 (those born 1930 or later) (Fig 1). For this study, we included everyone who had participated by January 2024. For the NONAGINTA study, 187 people had participated in the postal questionnaire in 2020-2023 (27%), and of them 96 also participated in the telephone interviews (13 full twin pairs; 8 MZ, DZ). Of them, 93 participants also had baseline questionnaire data from 1975.

**Fig 1.**
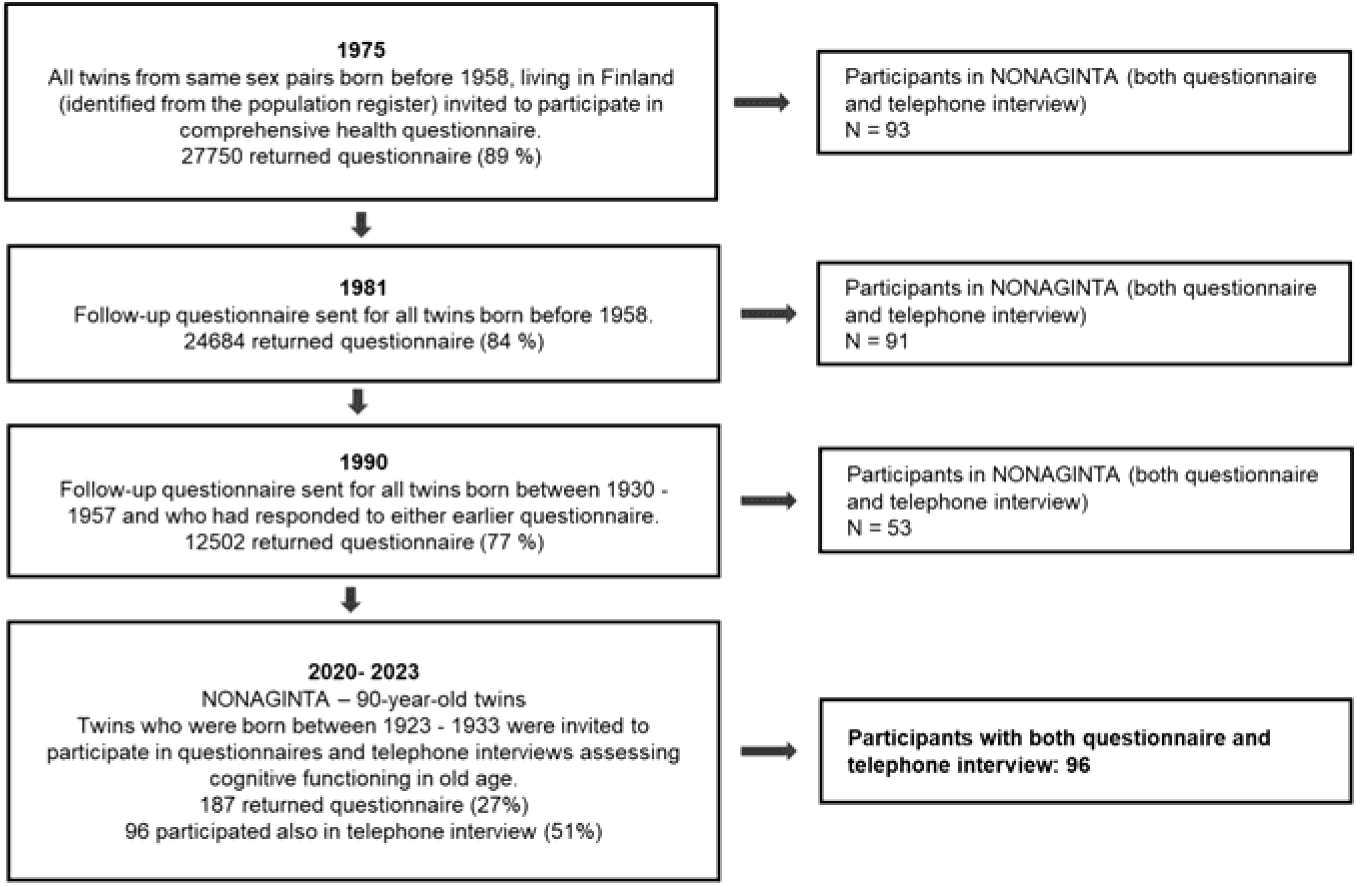
Flow chart of the study.

### Cardiovascular risk factors

The CV risk factors included BMI, BP, cholesterol, and PA. We also included education. These were self-reported measures assessed with a postal questionnaire at midlife in 1975 (baseline), 1981, 1990 and 2020-2023 (≥ 90-years-old). The detailed descriptions of the CV risk factors and education are given in the Supplement.

BMI was calculated for 1975, 1981, 1990, and 2020-2023 based on self-reported weight and height (kg/m2) at the time of the questionnaire. We used mean BMI in 1975 and 1981, BMI in 1990, and at 90 years old in the analyses. In case of missing data from either 1975 or 1981 questionnaire, we used single value from the other questionnaire.

We used a binary BP variable, with high and normal BP groups, assessed in 1975, 1981, 1990 and 2020-2023. In 1975, 3 of the 6 participants and in 1981, 5 of the 13 participants reporting high BP used anti-hypertensive medications, respectively. Of those with normal BP, only one person out of 82 have used anti-hypertensives briefly in 1975, while none in 1981.

A history of total cholesterol level was assessed in 1981, 1990, and 2020-2023 questionnaires (not included in 1975). Total cholesterol level was also transformed into a binary variable, with normal and high cholesterol groups. One person reported low cholesterol, and was categorized into the normal cholesterol group.

PA was assessed as MET (metabolic equivalent of task) hours expended per day (continuous variable) [14]. We used the mean of 1975 and 1981 MET hours/day, MET hours/day measured in 1990, and MET hours/day measured at 90 years old in the analyses. In case of missing data from either 1975 or 1981 questionnaire, we used single value from the other questionnaire.

For descriptive purposes we used self-reported measures of the presence of doctor-diagnosed diabetes (yes/no) [8], frequency of alcohol use (see the Supplement, S1 Table) and smoking status (yes/no) [15], in 1975, 1981, 1990, and at 90 years old.

### Education

Education was assessed with a self-reported measure in the questionnaires in 1975, 1981, and 2020-2023. For this study, we used a three-category education variable (6 years or less, 7-11 years, and 12 years or more). In the analyses, in each time point (1975, 1981, 1990, and 2020-2023) we used the education variable that was reported in the same timepoint as the risk factors were measured. For 1990, education from 1981 was used. Missing values were filled in based on most recent questionnaire.

### CAIDE

CAIDE score was based on [8], and consisted of midlife age, and self-report education, sex, BP, BMI, cholesterol, and PA (as opposed to original CAIDE with in-person measurements [11]). The CAIDE score ranges from 0-15, with higher values indicating higher dementia risk. See detailed description in the Supplement. Explanation on individual risk factors transformed into a total CAIDE score can be found in [8].

### Educational-occupational score

The educational-occupational score was based on [12]. It consisted of the following self-reported variables: age, years of education, work status, complexity of work, physical loading of work and work environment [8]. The questions for these variables and their categorization are explained in detail in [8].

### Cognitive measures

Three cognitive measures were used from a telephone interview-based cognitive assessment at age 90 years. Semantic fluency was measured with one-minute animal naming. Episodic memory was assessed with two measures including immediate and delayed recall of 10-word list from the modified Telephone Interview for Cognitive Status [16,17] using modification including three learning trials [18]. Immediate recall measure was total numbers of words recalled in trials 1-3. In delayed recall, participants were asked to repeat the words again after about 3 min from the immediate recall. The number of animals named, and words recalled in trials 1-3 and after delay were used as dependent variables for semantic fluency, immediate, and delayed recall, respectively.

### Statistical analyses

We used linear regression analyses with semantic fluency and immediate recall as dependent variables and individual CV risk factors, education, total CAIDE score, CAIDE without education, and educational-occupational score as independent variables. For the delayed recall, we used negative binomial regression due to distribution skewness towards zero values. We ran individual-level analyses (between-family) and adjusted for the non-independent data structure (family data). Regression analyses were conducted for each CV risk factor separately with three sets of covariates. Model 1 included age, sex, and follow-up time as covariates, model 2 included education as an additional covariate to model 1 covariates, and model 3 included *APOE* genotype as additional covariate to model 2 covariates. For *APOE* status, participants were categorized into ε4-carriers and non-carriers (more detail in the Supplement).

We conducted drop-out analyses (design-based F-test and two-sample t-test accounting for family relatedness) comparing midlife and old age risk factors and dementia risk scores between different participation groups: those who completed both a telephone interview and postal questionnaire at age 90, those who completed only postal questionnaire, and those who were invited but did not participate. All analyses were run on Stata MP 18 (64-bit).

### Ethical considerations

The NONAGINTA study was approved by the Coordinating Ethics Committee of the Hospital District of Helsinki and Uusimaa (HUS). The participants gave written informed consent for their participation. Returning questionnaires in 1975, 1981, and 1990 were considered as consent to participate and consistent with Finnish legislation on medical research at that time.

## Results

### Descriptive statistics

The descriptive characteristics are described in Table 1. The average follow-up time from baseline to NONAGINTA study was 45.92 years. Mean age was 45 years (standard deviation (SD)=2.27) at the time of the 1975 questionnaire, and 91 (SD=1.93) for questionnaire sent in 2020-2023.

**Table 1.** Descriptive statistics for those who participated in telephone interviews and questionnaires in 90-year-old data collection.

| **Characteristics** | **N** | **All**<br>**(96)** | **Men**<br>**(41)** | **Women**<br>**(55)** |
| --- | --- | --- | --- | --- |
| **All (90 years old)** |  | Mean (SD) | Mean (SD) | Mean (SD) |
| **Age (years)** |  |  |  |  |
| 1975 | 92 | 45.42 (2.27) | 45.84 (2.33) | 45.13 (2.21) |
| 1981 | 90 | 51.80 (2.43) | 52.08 (2.26) | 51.59 (2.23) |
| 1990 | 53 | 59.27 (1.11) | 59.31 (1.11) | 59.25 (1.12) |
| 2020-2023 | 96 | 91.22 (1.93) | 91.27 (1.95) | 91.20 (1.93) |
| **BMI (kg/m<sup>2</sup>)** |  |  |  |  |
| 1975/1981 | 94 | 24.67 (3.05) | 24.77 (2.46) | 24.60 (3.44) |
| 1990 | 51 | 25.78 (3.72) | 25.43 (3.09) | 25.97 (4.06) |
| 2020-2023 | 95 | 25.15 (4.10) | 24.56 (3.50) | 25.60 (4.48) |
| **Blood pressure (high/normal)** |  |  |  |  |
| 1975 | 91 | 6/85 | 2/36 | 4/49 |
| 1981 | 85 | 13/72 | 4/34 | 9/38 |
| 1990 | 49 | 11/38 | 3/14 | 8/24 |
| 2020-2023 | 90 | 63/27 | 22/17 | 41/10 |
| **Cholesterol (high/normal)** |  |  |  |  |
| 1981 | 36 | 7/29 | 5/18 | 2/11 |
| 1990 | 42 | 23/19 | 6/10 | 17/9 |
| 2020-2023 | 65 | 31/34 | 14/17 | 17/17 |
| **Physical activity (MET hours/day)** |  |  |  |  |
| 1975/1981 | 88 | 2.40 (1.71) | 2.74 (1.85) | 2.14 (1.57) |
| 1990 | 52 | 3.24 (4.56) | 4.88 (7.15) | 2.36 (1.87) |
| 2020-2023 | 84 | 1.70 (1.65) | 1.79 (1.48) | 1.64 (1.79) |
| **CAIDE total** | 54 | 7.61 (1.92) | 7.57 (1.73) | 7.65 (2.13) |
| **EDU-OCU** | 94 | 16.77 (3.15) | 16.68 (3.28) | 16.83 (3.08) |
| **EDU 1975** (<6yrs/ 7-11 yrs./ >12 yrs.) | 82 | 33/40/19 | 11/19/8 | 22/21/11 |
| **EDU 1981** (<6yrs/ 7-11 yrs./ >12 yrs.) | 94 | 33/41/20 | 13/18/9 | 20/23/11 |
| **EDU 90 yrs.** (<6yrs/ 7-11 yrs./ >12 yrs.) | 96 | 38/38/20 | 17/15/9 | 21/23/11 |
| **Semantic fluency** | 96 | 15.20 (4.92) | 15.73 (4.88) | 14.80 (4.95) |
| **Immediate recall** | 96 | 11.34 (4.45) | 11.02 (4.23) | 11.58 (4.63) |
| **Delayed recall** | 96 | 2.21 (2.27) | 1.88 (1.76) | 2.45 (2.57) |
| **APOE status** ( ε4 carrier/non-carrier) | 83 | 20/65 | 5/31 | 15/34 |

*APOE* = apolipoprotein E, BMI = body mass index, CAIDE = Cardiovascular Risk Factors, Aging and Dementia score, EDU = education, EDU-OCU = educational-occupational score, MET = metabolic equivalent hours per day, SD = standard deviation, yrs. = years.

The average number of words in semantic fluency was 15.20 (SD=4.92), 11.34 (SD=4.45) for immediate recall, and 2.21 (SD=2.27) for delayed recall. The participants had 8.97 (SD=4.45) years of education on average. A total of 19 individuals were *APOE* ε4-carriers (22%).

For BP, 6% (6/91) reported having high BP in 1975, 15% (13/85) in 1981, and 22% (11/49) in 1990. In the 2020-2023 questionnaire, 70% (63/ 90) reported having high BP. For cholesterol, 19% (7/36) reported having high cholesterol in midlife (1981), 55% (23/42) in late midlife (1990) and 48% (31/65) in old age (2020-2023). For midlife PA, mean MET hours/day were 2.40 (SD=1.71), for late midlife 3.24 (SD=4.56), and in old age, the mean was 1.70 (SD=1.65). The mean midlife BMI was 24.67 (SD=3.05), late midlife was on average 25.78 (SD=3.72), and in old age the mean was 25.15 (SD=4.10). The mean total CAIDE score was 7.61 (SD=1.92) and the mean educational-occupational score was 16.77 (SD=3.15).

In 1975 there were 11 current smokers, but only three in 1990 and as 90-year-olds. Nobody reported diabetes in 1975, 1981 or 1990 but among the 90+ year olds, 12 had type 2 diabetes. Most participants reported very little or no use of alcohol (S1 Table).

## Individual protective and risk factors

### Education

All results for risk and protective factors and dementia risk scores are displayed in Fig 2 (semantic fluency), Fig 3 (immediate recall), and Fig 4 (delayed recall). For semantic fluency, those with 12 or more years of education scored higher than those with 6 years or less (b=4.48, 95%CI: 1.50; 7.44, p=0.004) (reference group) and the result was similar when controlling for *APOE* (S2 Table). Those with 7-11 years of education (b=2.63, 95%CI: 0.58; 4.68, p=0.013) and those with 12 or more years of education (b=5.36, 95%CI: 3.75; 6.98, p<0.001) remembered more words in immediate recall compared to those with 6 years or less education. The results were similar when including *APOE* as a covariate (S2 Table). For delayed recall, those with 7-11 years of education and those with 12 or more years of education scored higher compared to those with 6 years or less education: b=0.49, 95%CI: - 0.01; 0.99, p=0.06, and b=1.11, 95%CI: 0.75; 1.46, p<0.001. The effect size remained essentially the same for 12 or more years of education when including *APOE* as covariate (b=1.04, 95%CI: 0.63; 1.45, p<0.001) (S2 Table).

**Fig 2.**
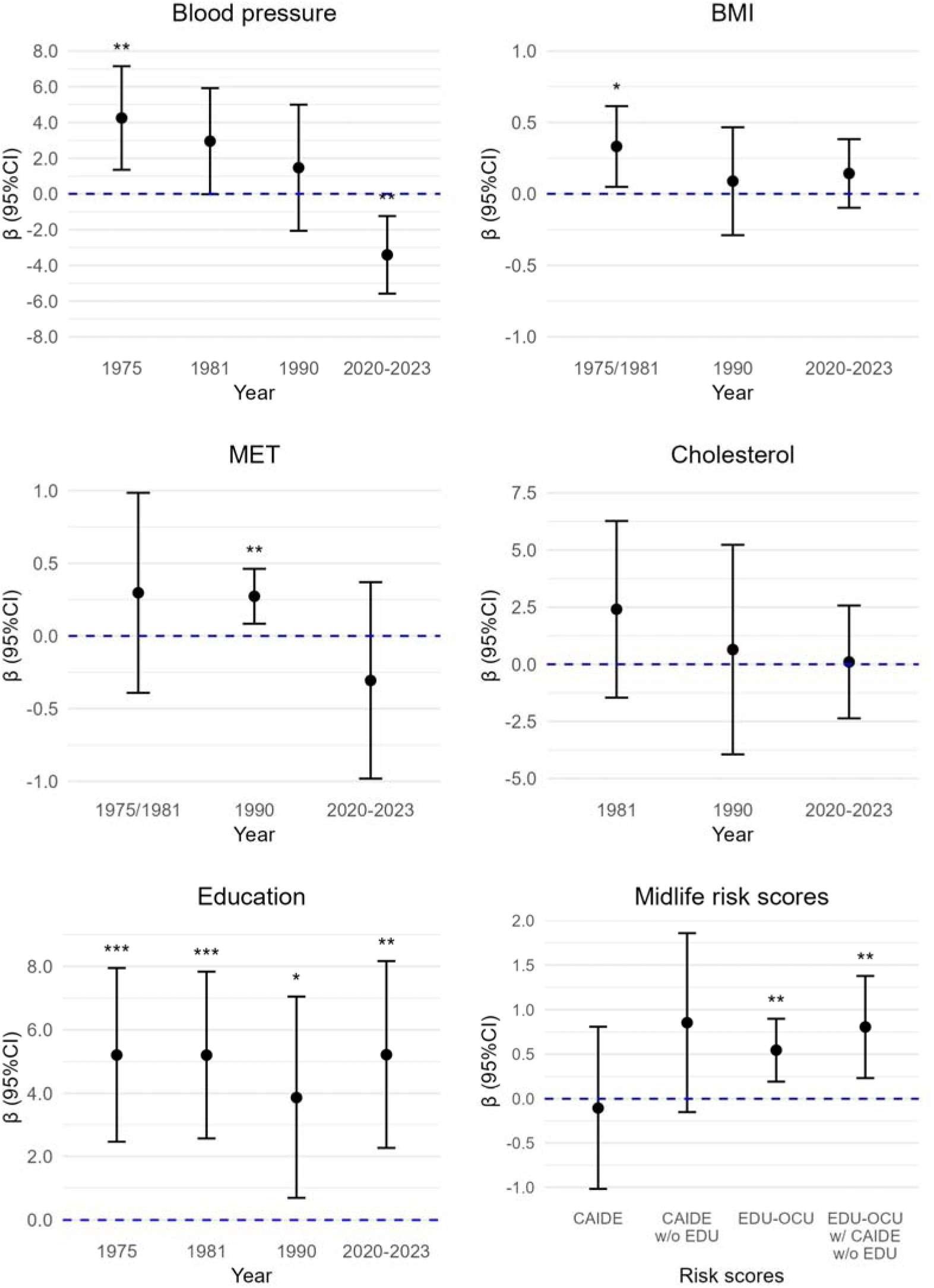
Cardiovascular risk factors and midlife risk scores in association with semantic fluency. BMI = body mass index, CAIDE = Cardiovascular Risk Factors, Aging and Dementia score, CI = confidence intervals, EDU = education, EDU-OCU = educational-occupational score, MET = metabolic equivalent hours per day, w/o = without. *p < 0.05. **p < 0.01. ***p < 0.001.

**Fig 3.**
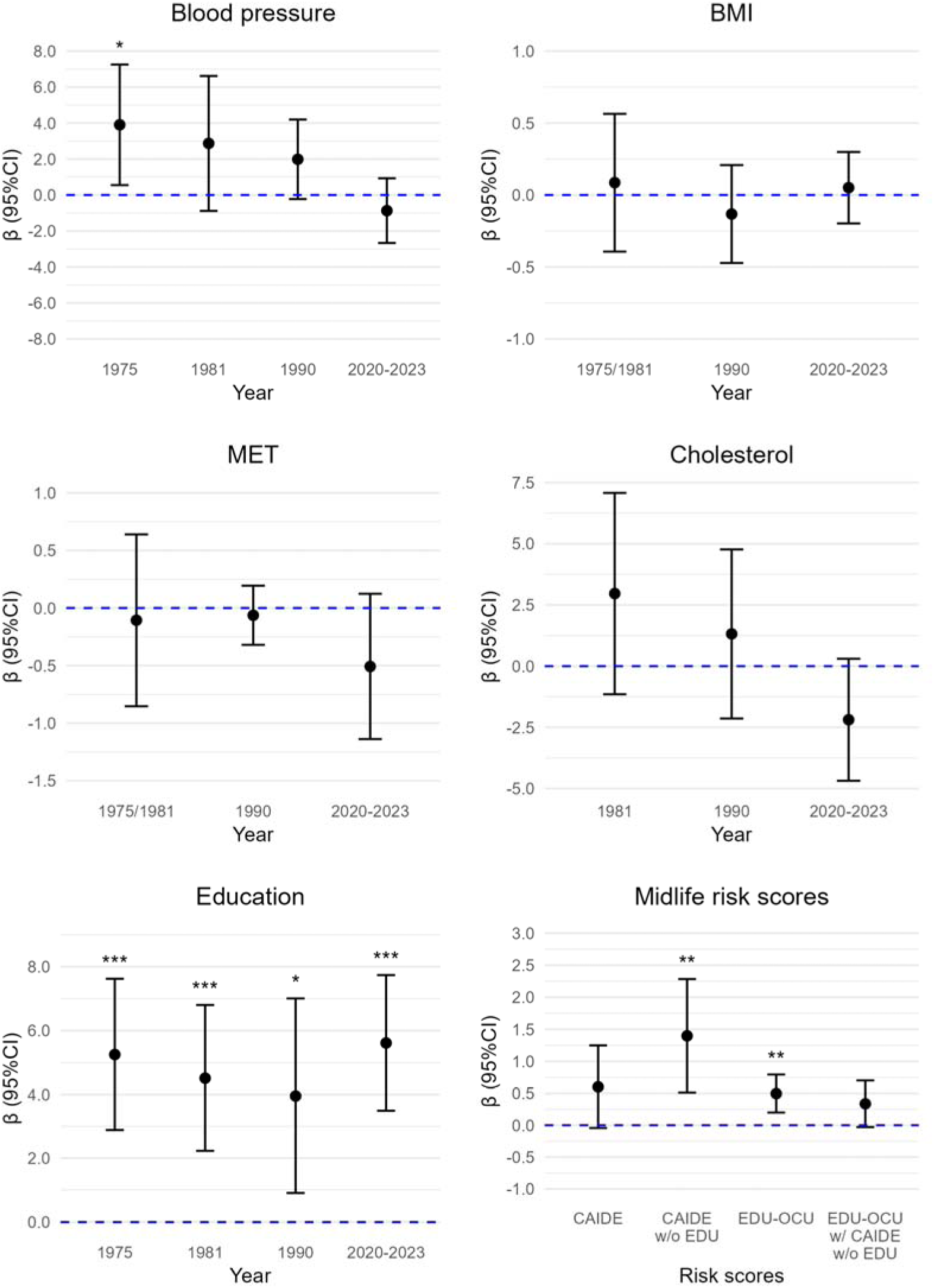
Cardiovascular risk factors and midlife risk scores in association with immediate recall. BMI = body mass index, CAIDE = Cardiovascular Risk Factors, Aging and Dementia score, CI = confidence intervals, EDU = education, EDU-OCU = educational-occupational score, MET = metabolic equivalent hours per day, w/o = without. *p < 0.05. **p < 0.01. ***p < 0.001.

**Fig 4.**
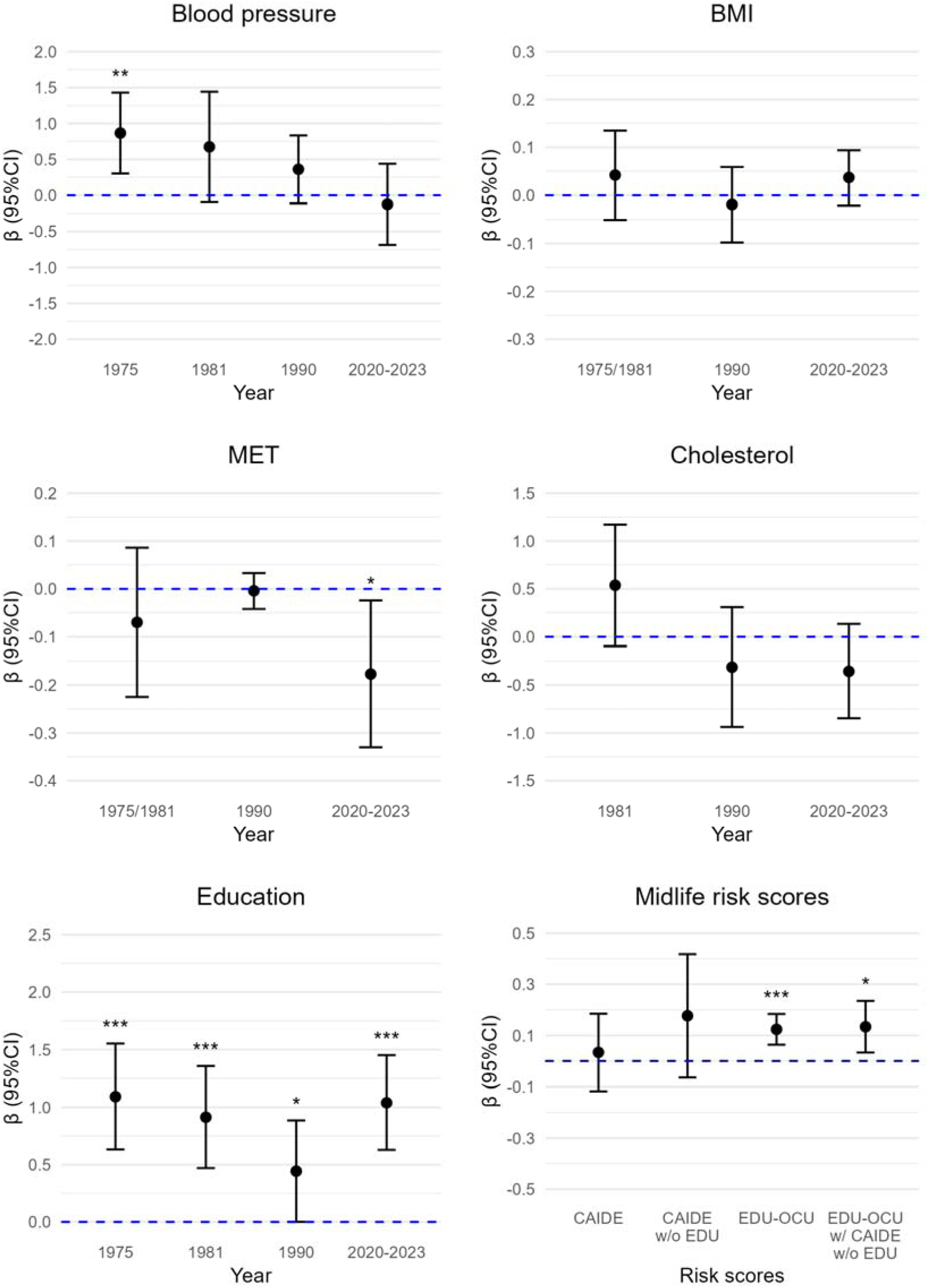
Cardiovascular risk factors and midlife risk scores in association with delayed recall. BMI = body mass index, CAIDE = Cardiovascular Risk Factors, Aging and Dementia score, CI = confidence intervals, EDU = education, EDU-OCU = educational-occupational score, MET = metabolic equivalent hours per day, w/o = without. *p < 0.05. **p < 0.01. ***p < 0.001.

### CV risk factors

Higher midlife BMI was associated with better semantic fluency (b=0.29, 95%CI: 0.002; 0.58, p= 0.05), also with *APOE* as covariate (S3 Table). Midlife BMI was not significantly associated with immediate recall or delayed recall (S3 Table). BMI in late midlife was not associated with any of the cognitive measures (S4 Table). Old age BMI was not associated with any cognitive measures (S2 Table).

Midlife PA was not significantly related to any of the cognitive measures (S3 Table). A higher PA in late midlife was associated with better semantic fluency (b=0.22, 95%CI: 0.04; 0.34, p= 0.02), also when accounting for *APOE*, but not with immediate or delayed recall (S4 Table). Old age PA was also not associated with semantic fluency or immediate recall (S2 Table). Higher old age PA was associated with lower scores in delayed recall (b=-.15, 95%CI:-0.28;-0.02, p= 0.02), also with *APOE* in the model (S2 Table).

Those who reported high BP in 1975 scored higher in semantic fluency (b=3.10, 95%CI: 0.43; 5.77, p= 0.02), immediate recall (b=3.27, 95%CI:0.33; 6.21, p=0.03) and delayed recall (b=0.87, 95%CI: 0.41; 1.32, p< 0.001) compared to those with normal BP, also with *APOE* in the model (S5 Table). People with high BP in 1981 showed better semantic fluency: (b=2.97, 95%CI: 0.22; 5.73, p= 0.04), also with *APOE* as a covariate. There was no significant association for 1981 BP in immediate recall, but people with high BP scored higher on delayed recall (b=0.68, 95%CI: 0.04; 1.32, p= 0.04) (S3 Table). This was not statistically significant when including *APOE* as a covariate. BP in late midlife was not associated with any of the cognitive measures (S4 Table). Those who reported high BP in old age scored lower in semantic fluency (b=-2.89, 95%CI:-4.86;-0.91, p= 0.005), and with *APOE* in the model, but this result was not significant in immediate recall or delayed recall (S2 Table).

Total cholesterol level at any age was not significantly related to any cognitive measures (we had no measure for cholesterol in 1975) (S2-S4 Tables).

## Midlife dementia risk scores

### CAIDE

Total CAIDE score was not significantly associated with semantic fluency, immediate recall, or delayed recall and the results were similar with or without *APOE* as a covariate (S6 Table). Higher CAIDE score without education was associated with better semantic fluency (b=1.05, 95%CI: 0.05; 2.05, p=0.04), but not with *APOE* in the model. Higher CAIDE score without education was also associated with better immediate recall (b=1.32, 95%CI: 0.44; 2.19, p=0.004) also with *APOE* in the model, but not with delayed recall, with or without *APOE* (S6 Table).

### Educational-occupational score

Educational-occupational score was positively associated with semantic fluency (b=0.47, 95%CI: 0.13; 0.82, p=0.008) even after adjusting for *APOE* (S6 Table). The results were similar with CAIDE risk score in the model. Educational-occupational score was also positively associated with immediate recall (b=0.54, 95%CI: 0.31; 0.78, p<0.001), and with delayed recall (b=0.13, 95%CI: 0.08; 0.18, p<0.001), and the results were similar with CAIDE score or *APOE* in the model (S6 Table).

### Co-twin analyses

This study included 13 full twin pairs (8 MZ and 5 DZ pairs). There were only few twin pairs discordant for CV risk factors, in midlife or old age (S7-S13 Tables). Detailed description of discordancy can be found in the Supplement.

### Drop-out analyses

Participants who completed both the telephone interview and questionnaire at age 90 had significantly higher educational-occupational scores (p<0.001), lower total CAIDE scores (p=0.006), and lower CAIDE score without education (p=0.029) than those who completed only the questionnaire. They also had higher educational-occupational scores (p<0.001) and lower CAIDE scores (p=0.013) than those who did not participate in neither. Participants were less likely to have high BP in 1990 than those with only questionnaires (p=0.012). They also had higher education levels in 1975, 1981, and at age 90 (all p<0.001). See Supplement for detailed results.

## Discussion

In line with previous studies on younger cohorts [7,19] and in 90-year olds [20], higher education was associated with better cognitive functioning at 90 years, suggesting that education protects cognition across the life span. This link could be due to people with higher education having more cognitively challenging jobs [21], or having higher general cognitive ability [22]. In line with this, the educational-occupational score showed a positive association with cognition.

The individual CV risk factors in our study were included in the latest Lancet commission report on dementia risk factors [7]. While better overall CV health has been associated with decreased dementia risk [23], studies have found that modifiable risk factors impact cognition differently across life stages [3–6]. Some studies looking at CV risk factors measured in old age have found protective relationships with cognition, such as hypertension and BMI [6,9]. In our study, midlife or late-life PA, BMI, cholesterol, and CAIDE scores showed no link to cognition at age 90, suggesting CV risk factors may play a different role in cognitive health for those reaching very old age. It is also possible that cognitive risk factors after 90 differ from those affecting younger older adults (60+), as previous studies suggest [20].

People who belonged in the high BP group in 1975 and 1981 had better cognition in old age, showing quite large effect sizes. Midlife hypertension is one of the most prevalent dementia risk factors, with nearly 60% increased risk of all cause dementia [21]. Mid-to-late-life antihypertensive medication use has been linked to lower dementia risk, while untreated hypertension raises risk [24,25]. In our study, people who reported using antihypertensive medications in midlife belonged in the high BP group. The earlier start of antihypertensive medications could explain the unexpected association between high BP in midlife and better cognition in old age. In 1970s Finland, many were unaware of having high BP [26], and some participants may have been misclassified as having normal BP, while those on medication might represent a group with better service access.

This study’s strengths include a 48-year follow-up and a population-based sample with a nearly equal gender balance. The small sample and the high number of covariates in regression analyses should be considered when interpreting results. Participants had higher education and lower CAIDE scores than those who only completed the questionnaire at age 90 or did not participate. Plans to compare co-twins with differing risk factors were not conducted due to high concordance in risk factors and small number of twin pairs. Prior studies in Finland have shown good agreement between self-reports, medical records, and in person measures [27–29]. Lastly, 90-year-olds tend to have a high prevalence of risk factors (e.g., high BP, low PA), diluting differences between individuals [30]. Those who reach age 90 and older may also be a selected group with generally healthy lifestyle.

## Conclusion

Education remains as a protective factor for cognition in above 90-year-olds. Our results suggest that CV risk factors are of at most modest importance, but due to sample size and selection effects, more research is needed to confirm this. Access to education in early life is crucial in preventing cognitive decline and in decreasing dementia risk all the way to late old age.

## Data availability

Due to the consent given by the participants and easier identification of twin data, data is not publicly available. Data are available through the Institute for Molecular Medicine Finland (FIMM) Data Access Committee (DAC) for those with authorization, who have IRB/ethics approval and an institutionally approved study plan. Please contact the FIMM DAC for more details.

## Data Availability

Due to the consent given by the participants and easier identification of twin data, data is not publicly available. All data in the present study are available through the Institute for Molecular Medicine Finland (FIMM) Data Access Committee (DAC) for those with authorization, who have IRB/ethics approval and an institutionally approved study plan. Please contact the FIMM DAC for more details.

## Acknowledgements

We would like to thank all the twins who participated in the study.

## Notes

### Competing Interest Statement

The authors have declared no competing interest.

### Funding Statement

This work was supported by the Finnish Brain Foundation [to A.V]; funding from the University of Helsinki Doctoral Programme in Population Health [to A.V]; Orion Research Foundation [to P.I.M]; The Biomedicum Helsinki Foundation [to P.I.M]; Juho Vainio Foundation [to S.A]; Academy of Finland Center of Excellence in Complex Disease Genetics [grant number 352792 to J.K]; NONAGINTA data collection was funded by the Research Council of Finland [grant numbers 320109, 345988 to E.V]; the Research Council of Finland, [grant number 314639 to E.V], and the Sigrid Juselius Foundation Senior Fellowship [to E.V]. The funders played no role in the design, execution, analysis, or interpretation of data, or writing of the study.

### Author Declarations

The Coordinating Ethics Committee of the Hospital District of Helsinki and Uusimaa (HUS) gave ethical approval for the NONAGINTA study. The participants gave written informed consent for their participation. Returning questionnaires in 1975, 1981, and 1990 were considered as consent to participate and consistent with Finnish legislation on medical research at that time.

