## Supplementary material for "Midlife and old-age cardiovascular risk factors, educational attainment, and cognition at 90-years – population-based study with 48-years of follow-up"

### Education

Education was assessed with a self-reported measure in the questionnaires in 1975, 1981, and 2020-2023. The participants were asked to classify themselves by length of schooling, according to eight educational categories. The answers were then transformed into eight categories in the following way, each category corresponding to years of education: less than primary school (mean 3 years), primary school (6 years of education), at least 1 year of education such as vocational training in addition to primary school (7 years), junior high school (9 years), at least 1 year of education such as vocational training in addition to junior high school (10 years), high school graduate (12 years), at least 1 year of education such as vocational training in addition to high school (13 years), and university degree (18 years), some other schooling, what? If the participant had answered option nine (some other schooling, what?) and written an explanation, the explanation was investigated and if the level of education was clear that level was implemented. If the level was still not clear, it was treated as missing. Missing values were filled in based on the education that was reported most recently in the previous questionnaires. For this study, we used a three-category education variable, which was based on the eight-level categorical variable described above. The three categories included 6 years or less education, 7-11 years of education, and 12 or more years of education. In the analyses, in each time point (1975, 1981, 1990, and 2020-2023) we used the education variable that was reported in the same timepoint as the risk factors were measured. For analyses on the 1975 risk factors, we used the education measured in 1975. For 1981 risk factors, we used the education that was measured in 1981, and filled in any missing values based on the 1975 education. In 1990 risk factors, we used the 1981 education and filled in missing values from 1975 (education was not measured in 1990). For mean body mass index (BMI) and physical activity (PA) (from 1975/1981) analysis we used education from 1981 and filled in missing values from 1975. For analyses on the 2020-2023 risk factors, we used the education that was measured at 90 years old, and filled in any missing values from the most recent questionnaire (1981 education, and if that was missing, we used education from 1975). There were six cases in which education measured at 90 years was reported lower than in 1975, and six cases in which education at 90-year-old was lower than in 1981.

### Blood pressure

Blood pressure (BP) was assessed with the following question in 1975: Has a doctor ever told you that you have or have had high BP? Answer options were yes/no. In 1981, 1990, and 2020-2023 questionnaires, the BP question was the following: Has a nurse or a doctor measured your BP within the last five years? For those who answered yes, the next question had four answer options: 1) BP was found to be normal, 2) BP was mildly raised, no medication, 3) raised and been on medication that has now ended, and 4) permanently raised with continuous medication. We used a binary BP variable, with high BP and normal BP groups. Those who answered “BP was found to be normal” were categorized into the normal BP group, and all other answer options belonged to the high BP group.

In 1975 and 1981 questionnaires, the use of antihypertensive medication was assessed with a question “How many days in the past year have you used the following medications? (including antihypertensive medications). Answer options were: 1) I have not used at all, 2) less than 10 days, 3) 10-59 days 4) 60-180 days (2-6 months), 5) over 180 days (over 6 months). In 2020-2023 questionnaire, the participants were asked to list all medications they were using.

### Cholesterol

Serum total cholesterol level was assessed with the following question in 1981, 1990, and 2020-2023 (not included in 1975): Has a nurse or a doctor measured your cholesterol within the last five years? For those who answered yes, the next question had three answer options: 1) found to be low, 2) found to be normal, and 3) found to be high. Cholesterol was also transformed into a binary variable, with normal cholesterol and high cholesterol groups. One person reported low cholesterol, and was categorized into the normal cholesterol group.

### Physical activity

Physical activity (PA) was assessed as MET (metabolic equivalent of task) hours expended per day (continuous variable). The PA questions that were used to calculate the MET hours/day included four questions assessing intensity, duration, frequency, and PA related to getting to and from work. The question about intensity (“The PA in your leisure time usually corresponds to”) had four answer options: 1) walking, 2) vigorous walking to jogging, 3) jogging, and 4) running. The question about duration (“How long does one leisure time PA session last on average?”) had five answer options: 1) less than 15 minutes, 2) 15 minutes or more but less than 30 minutes, 3) 30 minutes or more but less than an hour, 4) an hour or more but less than two hours, and 5) over two hours. The frequency question (“How many times a month do you engage in leisure time PA?”) included six answer options: 1) less than once a month, 2) one to two times a month, 3) three to five times a month, 4) six to ten times a month, 5) 11-19 times a month, and 6) over 20 times a month. The question regarding PA to and from work (“Daily I spent time commuting while engaging in walking, biking, running, and/or cross country skiing”) included five answer options: 1) less than 15 minutes, 2) 15 minutes or more but less than 30 minutes, 3) 30 minutes or more but less than an hour, 4) an hour or longer, and 5) currently I am not working. MET hours/day were calculated as the product of intensity*duration*frequency of PA. In the calculation, we used the following MET values for the intensity of PA: 4 (for PA intensity corresponding to walking), 6 (vigorous walking to jogging), 10 (jogging), and 13 (running) (1). The values of MET hours/day represent leisure-time PA because work-related PA was excluded, except at age 90 years when participants were retired and did not work anymore. We used the mean of 1975 and 1981 MET hours/day, MET hours/day measured in 1990, and MET hours/day measured at 90 years old in the analyses. In case of missing data from either 1975 or 1981 questionnaire, we used single value from the other questionnaire.

### CAIDE

Cardiovascular Risk Factors, Aging and Dementia (CAIDE) score was based on (2), and consisted of age, education, sex, BP, BMI, cholesterol, and PA. This CAIDE score was calculated based on self-reported measures, as opposed to original CAIDE with in-person measurements (3). Highest education from 1975 or 1981 was used. BP from 1981 was used, or from 1975 if missing in 1981. Mean BMI of 1975 and 1981 was used, or a single time point in case one was missing. Cholesterol was based on self-reported levels from 1981, or in case of missing data, the information from 1990 questionnaire was used. PA was based on 1981 questionnaire, and in case of missing information, data from 1975 was used. We also included a CAIDE score without education in the analyses. The CAIDE score ranges from 0-15, with higher values indicating higher dementia risk. Detailed explanation on how the individual risk factors were transformed into a total CAIDE score can be found in (2).

### Educational-occupational score

The educational-occupational score was based on earlier work as described by (4). It consisted of the following self-reported variables: age, years of education, work status, complexity of work, physical loading of work and work environment (2). Education was determined as the highest reported years either from 1975 or 1981 questionnaire. For work status, complexity of work, physical loading of work and work environment, data from 1981 was used (if missing, data from 1975 was used). The educational-occupational score ranges from 0-25, with higher scores indicating higher educational-occupational attainment. The questions for these variables and their categorization are explained in detail in (2).

### Apolipoprotein *E*

Venous blood samples were collected from participants at local health centers using kits mailed to them in advance. The samples, along with signed consent forms, were then returned to the Finnish Institute for Health and Welfare (until 2008 called National Public Health Institute) in Helsinki, Finland, for Apolipoprotein *E* (*APOE*) genotyping. DNA was extracted and stored for subsequent analysis. For participants who had previously provided DNA samples, re-consent was obtained through telephone interviewers, followed by signed consent forms for this study. *APOE* genotyping was conducted by analyzing two single-nucleotide polymorphisms, rs429358 and rs7412 (5).

### Co-twin analyses

This study included 13 full twin pairs, with 8 MZ and 5 DZ pairs. Five out of 13 twin pairs were discordant for education in old age. At 90-years old, six out of 12 pairs were discordant for low vs. high BMI (cutoff 25), nine out of 10 for PA, six out of 12 for BP, and two out of five pairs for cholesterol. In midlife, five out of 13 pairs were discordant for BMI, two out of three for cholesterol, and 12 out of 13 were discordant for midlife PA. There was one twin pair (out of 13) discordant for BP in 1975. In 1981, there were no pairs discordant for BP (out of 12). For total CAIDE score, there were five discordant twin pairs (out of 6) and for educational occupational score, there were eight discordant pairs out of 13. Cross tables of discordant pairs for each risk factor and semantic fluency are shown in S-13 Tables (only risk factors with more than six discordant pairs are shown).

### Drop-out analyses

We conducted drop-out analysis comparing continuous variables (BMI, PA, total CAIDE score, CAIDE without education, and educational-occupational score) in midlife and old age between those who participated in both telephone interview and questionnaire at 90 years old and those who only participated in questionnaire in the 90-year-old data collection. We also compared those who participated in both telephone interview and questionnaire, and those who were alive and invited to participate in the questionnaire at 90 years old, but who did not participate in either one. We used two sample t-tests, accounting for family relatedness. Descriptive statistics for those who only participated in the questionnaires can be seen in S14 Table. Results for t-tests can be seen in S15 Table. Those who participated in telephone interview and questionnaire at 90 years had significantly higher educational-occupational scores (mean (M)=16.77, standard deviation (SD)=3.15) than those with only a questionnaire at 90 (M=15.08, SD=2.26; *t*(160)= -3.75, p<0.001, 95%CI: -2.58; -0.80). Those with telephone interview and questionnaire also had significantly lower total CAIDE scores (M= 7.61, SD=1.92) compared to those with only questionnaire at 90 years old (M=8.89, SD=2.22; *t*(87)=2.82, p=0.006, 95%CI: 0.38; 2.17). The results were similar for CAIDE scores without education, with those with telephone interviews having lower scores (M=5.96, SD=1.36) compared to those with only questionnaire (M=6.70, SD=1.84; *t*(87)=2.22, p=0.029, 95%CI: 0.08; 1.41).

Participants with both telephone interview and questionnaire had significantly higher educational-occupational scores (M=16.77, SD=3.15) than those who did not participate in either (M=14.91, SD=2.63; *t*(494)=-4.84, p<0.001, 95%CI: -2.61; -1.10). Those with telephone interviews had lower total CAIDE scores (M=7.61, SD=1.92) than those who did not participate in either (M=8.40, SD=1.93; *t*(239)=2.50, p=0.013, 95%CI: 0.17; 1.40).

A design-based F-test was conducted to examine the association between participation group (telephone interview and questionnaire, only 90-year questionnaire, or neither), and the categorical risk variables, including BP (high vs. normal), cholesterol (high vs. normal) and education (6 years or less, 7-11 years, and 12 years or more). F-test results are shown in S16 Table. There was a statistically significant association for 1990 BP and participation group of those with telephone interviews and those with only questionnaires (F(1, 79)= 6.70, p=0.012), with people with telephone interviews less likely to have high BP. Participants with telephone interviews also had higher education than those with only questionnaires: 1975 education (F(2, 307.73)=9.55, p<0.001), 1981 education (F(1.96, 314.95)=8.05, p<0.001) and education at 90 years old (F(1.94,318.05)=18.99, p<0.001).

For those with telephone interviews and questionnaires at 90 years, and those who did not participate in either one, there was a significant association between participation group and 1975 education (F(2.00, 914.60)=19.19, p<0.001) and 1981 education (F(2.00, 1005.83)=19.59, p<0.001), those with telephone interviews having higher education.

### Tables

**S1 Table. Alcohol use in a month. Those who participated in telephone interviews and questionnaires at 90 years old. Highest reported frequency of use for any category of alcoholic beverage (beer, wine, or liquor).**

|  | **N** | | | |
| --- | --- | --- | --- | --- |
| **How often do you use alcoholic beverages?** | **1975** | **1981** | **1990** | **90 yrs.** |
| Never | 22 | 21 | 10 | 40 |
| Less than two days a month on average | 38 | 47 | 25 | 32 |
| 3-8 days / month | 24 | 13 | 13 | 17 |
| 9-16 days / month | 6 | 9 | 3 | 2 |
| over 16 days / month | 2 | 1 | 1 | 5 |

yrs. = years.

**S2 Table. Linear regression analysis results for lifestyle factors at 90 years old predicting semantic fluency, immediate recall, and delayed recall at 90 years old.**

|  |  |  | **Semantic fluency** |  | **Immediate recall** |  | **Delayed recall** |  |
| --- | --- | --- | --- | --- | --- | --- | --- | --- |
|  | **Risk factor** | **N** | **b (95%CI)** | ***p*** | **b (95%CI)** | ***p*** | **b (95%CI)** | ***p*** |
| **Model 1** | BP | 90 | -3.01 (-5.24; -0.79) | 0.009 | -0.98 (-2.88; 0.93) | 0.312 | -0.13 (-0.59; 0.32) | 0.569 |
|  | Chol | 66 | 0.10 (-2.20; 2.41) | 0.929 | -2.34 (-4.69; 0.01) | 0.051 | -0.32 (-0.77; 0.13) | 0.159 |
|  | BMI | 95 | 0.07 (-0.19; 0.33) | 0.585 | 0 (-0.27; 0.27) | 0.999 | 0.03 (-0.03; 0.08) | 0.313 |
|  | MET | 84 | -0.09 (-0.68; 0.50) | 0.765 | -0.22 (-0.84; 0.39) | 0.477 | -0.08 (-0.22; 0.07) | 0.306 |
|  | Edu lev 1 | 96 | 0.83 (-1.26; 2.91) | 0.432 | 2.63 (0.58; 4.68) | 0.013 | 0.49 (-0.01; 0.99) | 0.056 |
|  | Edu lev 2 | 96 | 4.47 (1.50; 7.44) | 0.004 | 5.36 (3.75; 6.98) | <0.001 | 1.11 (0.75; 1.46) | <0.001 |
| **Model 2** | BP | 90 | -2.89 (-4.86; -0.91) | 0.005 | -0.74 (-2.43; 0.95) | 0.385 | -0.15 (-0.58; 0.28) | 0.484 |
|  | Chol | 66 | 0.41 (-1.79; 2.61) | 0.711 | -1.94 (-4.09; 0.21) | 0.076 | -0.29 (-0.70; 0.13) | 0.175 |
|  | BMI | 95 | 0.10 (-0.14; 0.34) | 0.399 | 0.04 (-0.20; 0.28) | 0.760 | 0.04 (-0.02; 0.09) | 0.197 |
|  | MET | 84 | -0.34 (-0.94; 0.26) | 0.265 | -0.53 (-1.09; 0.04) | 0.068 | -0.15 (-0.28; -0.02) | 0.020 |
| **Model 3** | BP | 77 | -3.41 (-5.58; -1.24) | 0.003 | -0.87 (-2.66; 0.93) | 0.339 | -0.12 (-0.69; 0.44) | 0.666 |
|  | Chol | 56 | 0.10 (-2.37; 2.57) | 0.934 | -2.19 (-4.68; 0.30) | 0.084 | -0.36 (-0.85; 0.13) | 0.154 |
|  | BMI | 82 | 0.14 (-0.10; 0.38) | 0.240 | 0.05 (-0.20; 0.30) | 0.681 | 0.04 (-0.02; 0.09) | 0.215 |
|  | MET | 71 | -0.31 (-0.98; 0.37) | 0.369 | -0.51 (-1.14; 0.12) | 0.113 | -0.18 (-0.33; -0.02) | 0.024 |
|  | Edu lev 1* | 83 | 0.96 (-1.28; 3.20) | 0.397 | 2.81 (0.65; 4.96) | 0.011 | 0.42 (-0.70; 0.93) | 0.111 |
|  | Edu lev 2* | 83 | 5.22 (2.27; 8.17) | 0.001 | 5.61 (3.49; 7.74) | <0.001 | 1.04 (0.63; 1.45) | <0.001 |

BMI = body mass index, BP = blood pressure, Chol = cholesterol, CI = confidence intervals, EDU lev 1 = education category 1 (7–11 years), EDU lev 2 = education category 2 (above 12 years), MET = metabolic equivalent hours per day. Model 1: Sex, and age (centered) are used as covariates. Model 2: Sex, age (centered), and education are used as covariates. Model 3: Sex, age (centered), education, and APOE are used as covariates. Analyses adjusted for non-independence of twin data. *Covariates for education in model 3 were sex, age (centered), follow-up time (centered), and APOE status.

**S3 Table. Linear regression analysis results for lifestyle factors in 1981 predicting semantic fluency, immediate recall, and delayed recall at 90 years old.**

|  |  |  | **Semantic fluency** |  | **Immediate recall** |  | **Delayed recall** |  |
| --- | --- | --- | --- | --- | --- | --- | --- | --- |
|  | **Risk factor** | **N** | **b (95%CI)** | ***p*** | **b (95%CI)** | ***p*** | **b (95%CI)** | ***p*** |
| **Model 1** | BP | 85 | 3.54 (0.58; 6.51) | 0.020 | 3.27 (0.06; 6.49) | 0.046 | 0.74 (0.17; 1.31) | 0.012 |
|  | Chol | 36 | 1.59 (-1.81; 4.99) | 0.346 | 2.52 (-0.96; 6.01) | 0.150 | 0.15 (-0.55; 85) | 0.674 |
|  | BMI | 93 | 0.19 (-0.10; 0.49) | 0.192 | -0.08 (-0.49; 0.33) | 0.702 | 0 (-0.08; 0.08) | 0.991 |
|  | MET | 86 | 0.43 (-0.28; 1.14) | 0.230 | 0.17 (-0.52; 0.86) | 0.616 | 0.01 (-0.11; 0.14) | 0.842 |
|  | Edu lev 1 | 91 | 0.70 (-1.39; 2.78) | 0.509 | 1.52 (-0.86; 3.89) | 0.208 | 0.24 (-0.35; 0.83) | 0.419 |
|  | Edu lev 2 | 91 | 4.19 (1.29; 7.08) | 0.005 | 4.77 (2.88; 6.66) | <0.001 | 0.99 (0.60; 1.39) | <0.001 |
| **Model 2** | BP | 85 | 2.94 (0.18; 5.70) | 0.037 | 2.61 (-0.68; 5.89) | 0.118 | 0.68 (0.05; 1.30) | 0.036 |
|  | Chol | 36 | 2.09 (-1.42; 5.60) | 0.233 | 3.32 (-0.98; 7.61) | 0.126 | 0.39 (-0.26; 1.04) | 0.236 |
|  | BMI | 93 | 0.29 (0.002; 0.58) | 0.049 | 0.03 (-0.39; 0.45) | 0.888 | 0.02 (-0.07; 0.11) | 0.596 |
|  | MET | 86 | 0.28 (-0.38; 0.95) | 0.400 | -0.06 (-0.75; 0.62) | 0.851 | -0.06 (-0.19; 0.07) | 0.356 |
| **Model 3** | BP | 76 | 2.95 (-0.02; 5.92) | 0.052 | 2.87 (-0.88; 6.62) | 0.131 | 0.67 (-0.10; 1.44) | 0.086 |
|  | Chol | 34 | 2.41 (-1.46; 6.27) | 0.213 | 2.96 (-1.15; 7.07) | 0.152 | 0.54 (-0.10; 1.17) | 0.097 |
|  | BMI | 83 | 0.33 (0.05; 0.61) | 0.022 | 0.09 (-0.39; 0.56) | 0.722 | 0.04 (-0.05; 0.14) | 0.376 |
|  | MET | 77 | 0.30 (-0.39; 0.99) | 0.392 | -0.11 (-0.85; 0.64) | 0.777 | -0.07 (-0.23; 0.09) | 0.380 |
|  | Edu lev 1* | 81 | 0.71 (-1.59; 3.02) | 0.540 | 1.51 (-1.04; 4.06) | 0.242 | 0.17 (-0.44; 0.77) | 0.591 |
|  | Edu lev 2* | 81 | 5.20 (2.57; 7.83) | <0.001 | 4.52 (2.23; 6.80) | <0.001 | 0.91 (0.47; 1.36) | <0.001 |

BMI = body mass index, BP = blood pressure, Chol = cholesterol, CI = confidence intervals, EDU lev 1 = education category 1 (7–11 years), EDU lev 2 = education category 2 (above 12 years), MET = metabolic equivalent hours per day. Model 1: Sex, age (centered) and follow-up time (centered) are used as covariates. Model 2: Sex, age (centered), follow-up time (centered), and education are used as covariates. Model 3: Sex, age (centered), follow-up time (centered), education, and APOE are used as covariates. BMI and MET variables are mean values based on 1975 and 1981 questionnaires. Analyses adjusted for non-independence of twin data. *Covariates for education in model 3 were sex, age (centered), follow-up time (centered), and APOE status.

**S4 Table. Linear regression analysis results for lifestyle factors in 1990 predicting semantic fluency, immediate recall, and delayed recall at 90 years old.**

|  |  |  | **Semantic fluency** |  | **Immediate recall** |  | **Delayed recall** |  |
| --- | --- | --- | --- | --- | --- | --- | --- | --- |
| **Model 1** | **Risk factor** | **N** | **b (95%CI)** | ***p*** | **b (95%CI)** | ***p*** | **b (95%CI)** | ***p*** |
|  | BMI | 51 | 0.01 (-0.29; 0.31) | 0.937 | -0.22 (-0.55; 0.10) | 0.174 | -0.04 (-0.11; 0.03) | 0.223 |
|  | BP | 49 | 1.63 (-1.77; 5.02) | 0.339 | 1.05 (-1.48; 3.58) | 0.405 | 0.22 (-0.25; 0.68) | 0.360 |
|  | Chol | 42 | 2.19 (-1.28; 5.67) | 0.209 | 0.98 (-1.56; 3.53) | 0.438 | -0.11 (-0.64; 0.43) | 0.700 |
|  | MET | 52 | 0.22 (0.05; 0.38) | 0.010 | -0.03 (-0.33; 0.28) | 0.860 | 0.01 (-0.03; 0.05) | 0.700 |
|  | Edu lev 1 | 53 | -0.21 (-3.27; 2.86) | 0.892 | 1.91 (-0.80; 4.62) | 0.163 | 0.07 (-0.46; 0.61) | 0.790 |
|  | Edu lev 2 | 53 | 2.71 (-1.09; 6.51) | 0.157 | 3.97 (1.56; 6.38) | 0.002 | 0.58 (0.19; 0.97) | 0.004 |
| **Model 2** | BMI | 51 | 0.06 (-0.29; 0.41) | 0.737 | -0.11 (-0.42; 0.20) | 0.489 | -0.04 (-0.11; 0.04) | 0.370 |
|  | BP | 49 | 1.46 (-1.53; 4.44) | 0.330 | 0.970 (-1.26; 3.25) | 0.377 | 0.17 (-0.30; 0.65) | 0.471 |
|  | Chol | 42 | 1.41 (-3.14; 5.96) | 0.534 | 1.14 (-1.78; 4.05) | 0.434 | -0.28 (-0.87; 0.32) | 0.361 |
|  | MET | 52 | 0.22 (0.04; 0.40) | 0.017 | -0.07 (-0.33; 0.20) | 0.604 | 0.003 (-0.03; 0.04) | 0.850 |
| **Model 3** | BMI | 45 | 0.09 (-0.29; 0.47) | 0.638 | -0.13 (-0.47; 0.21) | 0.435 | -0.02 (-0.10; 0.06) | 0.632 |
|  | BP | 44 | 1.464 (-2.067; 4.99) | 0.406 | 1.99 (-0.22; 4.20) | 0.077 | 0.36 (-0.11; 0.83) | 0.134 |
|  | Chol | 37 | 0.64 (-3.94; 5.23) | 0.777 | 1.32 (-2.14; 4.77) | 0.443 | -0.31 (-0.94; 0.31) | 0.325 |
|  | MET | 46 | 0.27 (0.08; 0.46) | 0.006 | -0.06 (-0.32; 0.19) | 0.620 | -0.004 (-0.04; 0.03) | 0.821 |
|  | Edu lev 1* | 47 | -0.64 (-3.94; 2.66) | 0.696 | 2.23 (-0.98; 5.45) | 0.168 | 0.01 (-0.57; 0.59) | 0.973 |
|  | Edu lev 2* | 47 | 3.87 (0.69; 7.05) | 0.019 | 3.96 (0.90; 7.01) | 0.012 | 0.44 (0.001; 0.89) | 0.049 |

BMI = body mass index, BP = blood pressure, Chol = cholesterol, CI = confidence intervals, EDU lev 1 = education category 1 (7–11 years), EDU lev 2 = education category 2 (above 12 years), MET = metabolic equivalent hours per day. Model 1: Sex, age (centered) and follow-up time (centered) are used as covariates. Model 2: Sex, age (centered), follow-up time (centered), and education are used as covariates. Model 3: Sex, age (centered), follow-up time (centered), education, and APOE are used as covariates. Analyses adjusted for non-independence of twin data. *Covariates for education in model 3 were sex, age (centered), follow-up time (centered), and APOE status.

**S5 Table. Linear regression analysis results for lifestyle factors from 1975 predicting semantic fluency, immediate recall, and delayed recall at 90 years old.**

|  |  |  | **Semantic fluency** |  | ***Immediate recall*** |  | ***Delayed recall*** |  |
| --- | --- | --- | --- | --- | --- | --- | --- | --- |
|  | **Risk factor** | **N** | **b (95%CI)** | ***p*** | **b (95%CI)** | ***p*** | **b (95%CI)** | ***p*** |
| **Model 1** | BP | 91 | 3.03 (0.93; 5.14) | 0.005 | 3.01 (-0.13; 6.14) | 0.060 | 0.73 (0.23; 1.24) | 0.005 |
|  | EDU lev 1 | 93 | 0.96 (-1.19; 3.11) | 0.378 | 2.97 (0.73; 5.21) | 0.010 | 0.56 (0.02; 1.11) | 0.042 |
|  | EDU lev 2 | 93 | 4.16 (1.17; 7.15) | 0.007 | 5.55 (3.63; 7.46) | <0.001 | 1.19 (0.77; 1.60) | <0.001 |
| **Model 2** | BP | 91 | 3.09 (0.43; 5.77) | 0.024 | 3.27 (0.33; 6.21) | 0.030 | 0.87 (0.41; 1.32) | <0.001 |
| **Model 3** | BP | 81 | 4.24 (1.34; 7.14) | 0.005 | 3.90 (0.55; 7.25) | 0.023 | 0.87 (0.30; 1.43) | 0.003 |
|  | EDU lev 1* | 83 | 0.93 (-1.35; 3.21) | 0.419 | 2.92 (0.56; 5.28) | 0.016 | 0.51 (-0.04; 1.06) | 0.067 |
|  | EDU lev 2* | 83 | 5.21 (2.47; 7.95) | 0.000 | 5.25 (2.89; 7.62) | <0.001 | 1.09 (0.63; 1.55) | <0.001 |

CI = confidence intervals. BP = blood pressure. EDU lev 1 = education category 1 (7-11 years), EDU lev 2 = education category 2 (above 12 years). Model 1: Sex, age (centered) and follow-up time (centered) are used as covariates. Model 2: Sex, age (centered), follow-up time (centered), and education are used as covariates. Model 3: Sex, age (centered), follow-up time (centered), education, and APOE are used as covariates. Analyses adjusted for non-independence of twin data. *Covariates for education in model 3 were sex, age (centered), follow-up time (centered), and APOE status.

**S6 Table. Linear regression analysis results for midlife dementia risk scores, CAIDE and educational-occupational score, predicting semantic fluency, immediate recall, and delayed recall at 90 years old.**

|  |  |  | **Semantic fluency** |  | **Immediate recall** |  | **Delayed recall** |  |
| --- | --- | --- | --- | --- | --- | --- | --- | --- |
|  | **Risk score** | **N** | **b (95%CI)** | ***p*** | **b (95%CI)** | ***p*** | **b (95%CI)** | ***p*** |
| **Model 1** | CAIDE total | 54 | -0.01 (-0.92; 0.89) | 0.975 | 0.42 (-0.18; 1.01) | 0.169 | -0.002 (-0.15; 0.14) | 0.977 |
|  | CAIDE (w/o EDU) | 54 | 1.05 (0.05; 2.05) | 0.040 | 1.32 (0.44; 2.19) | 0.004 | 0.16 (-0.07; 0.39) | 0.186 |
|  | EDU-OCU | 94 | 0.47 (0.13; 0.82) | 0.008 | 0.54 (0.31; 0.78) | <0.001 | 0.13 (0.08; 0.18) | <0.001 |
|  | EDU-OCU  (+ CAIDE w/o EDU) | 54 | 0.65 (0.18; 1.11) | 0.007 | 0.37 (0.10; 0.64) | 0.009 | 0.12 (0.05; 0.20) | 0.002 |
| **Model 2** | CAIDE total | 47 | -0.11 (-1.02; 0.81) | 0.817 | 0.60 (-0.04; 1.25) | 0.066 | 0.03 (-0.12; 0.19) | 0.661 |
|  | CAIDE (w/o EDU) | 47 | 0.85 (-0.15; 1.86) | 0.094 | 1.40 (0.51; 2.28) | 0.003 | 0.18 (-0.06; 0.42) | 0.150 |
|  | EDU-OCU | 83 | 0.55 (0.19; 0.90) | 0.003 | 0.50 (0.20; 0.80) | 0.001 | 0.12 (0.06; 0.18) | <0.001 |
|  | EDU-OCU  (+ CAIDE w/o EDU) | 47 | 0.81 (0.23; 1.38) | 0.007 | 0.34 (-0.03; 0.70) | 0.070 | 0.13 (0.03; 0.24) | 0.010 |

CAIDE = Cardiovascular Risk Factors, Aging and Dementia score, CI = confidence intervals, EDU = education, EDU-OCU = educational-occupational score, w/o = without. Model 1: Follow-up time (centered) used as a covariate. Model 2: Follow-up time (centered), and APOE are used as covariates. Analyses adjusted for non-independence of twin data.

**S7 Table. Discordant twin pairs for semantic fluency and midlife BMI (cutoff 25).**

|  | **Control (lower BMI)** | |
| --- | --- | --- |
| **Case (higher BMI)** | Poorer memory | Better memory |
| Poorer memory | - | 2 |
| Better memory | 5 | - |

BMI = body mass index.

**S8 Table. Discordant twin pairs for semantic fluency and late midlife BMI (cutoff 25).**

|  | **Control (lower BMI)** | |
| --- | --- | --- |
| **Case (higher BMI)** | Poorer memory | Better memory |
| Poorer memory | - | 2 |
| Better memory | 4 | - |

BMI = body mass index.

**S9 Table. Discordant twin pairs for semantic fluency and old age BMI (cutoff 25).**

|  | **Control (lower BMI)** | |
| --- | --- | --- |
| **Case (higher BMI)** | Poorer memory | Better memory |
| Poorer memory | - | 2 |
| Better memory | 5 | - |

BMI = body mass index.

**S10 Table. Discordant twin pairs for semantic fluency and midlife physical activity.**

|  | **Control (higher MET)** | |
| --- | --- | --- |
| **Case (lower MET)** | Poorer memory | Better memory |
| Poorer memory | - | 4 |
| Better memory | 8 | - |

MET = metabolic equivalent hours per day.

**S11 Table. Discordant twin pairs for semantic fluency and late midlife physical activity (1990).**

|  | **Control (higher MET)** | |
| --- | --- | --- |
| **Case (lower MET)** | Poorer memory | Better memory |
| Poorer memory | - | 3 |
| Better memory | 7 | - |

MET = metabolic equivalent hours per day.

**S12 Table. Discordant twin pairs for semantic fluency and old age physical activity.**

|  | **Control (higher MET)** | |
| --- | --- | --- |
| **Case (lower MET)** | Poorer memory | Better memory |
| Poorer memory | - | 5 |
| Better memory | 4 | - |

MET = metabolic equivalent hours per day.

**S13 Table. Discordant twin pairs for semantic fluency and educational-occupational score.**

|  | **Control (higher EDU-OCU)** | |
| --- | --- | --- |
| **Case (lower EDU-OCU)** | Poorer memory | Better memory |
| Poorer memory | - | 2 |
| Better memory | 6 | - |

EDU-OCU = educational occupational score.

**S14 Table. Descriptive statistics for those who participated in telephone interviews and questionnaires in 90 years old, and those who only participated in questionnaire at age 90.**

| **Characteristics** | **N** | | **All** | **Men** | **Women** |
| --- | --- | --- | --- | --- | --- |
| **All (90 yrs.)** |  | | **(96)** | **(41)** | **(55)** |
| **No telephone interview (90 yrs.)** |  | | **(91)** | **(21)** | **(70)** |
|  |  | | Mean (SD) | Mean (SD) | Mean (SD) |
| **Age** (years) |  | |  |  |  |
| 1975 | 92 | | 45.42 (2.27) | 45.84 (2.33) | 45.13 (2.21) |
| 1975 w/o telephone interview | 84 | | 44.74 (2.43) | 44.26 (1.69) | 44.88 (2.60) |
| 1981 | 90 | | 51.80 (2.43) | 52.08 (2.26) | 51.59 (2.23) |
| 1981 w/o telephone interview | 84 | | 51.05 (2.43) | 50.70 (1.89) | 51.15 (2.57) |
| 1990 | 53 | | 59.27 (1.11) | 59.31 (1.11) | 59.25 (1.12) |
| 1990 w/o telephone interview | 49 | | 58.74 (1.23) | 58.55 (1.45) | 58.80 (1.18) |
| 2020-2023 | 96 | | 91.22 (1.93) | 91.27 (1.95) | 91.20 (1.93) |
| 2020-2023 w/o telephone interview | 90 | | 90.98 (1.79) | 90.5 (0.95) | 91.11 (1.95) |
| **BMI** (kg/m^2^) |  | |  |  |  |
| 1975/1981 | 94 | | 24.67 (3.05) | 24.77 (2.46) | 24.60 (3.44) |
| 1975/1981 w/o telephone interview | 90 | | 24.22 (2.65) | 24.83 (2.26) | 24.04 (2.74) |
| 1990 | 51 | | 25.78 (3.72) | 25.43 (3.09) | 25.97 (4.06) |
| 1990 w/o telephone interview | 48 | | 25.28(2.96) | 25.57 (2.29) | 25.14 (3.21) |
| 2020-2023 | 95 | | 25.15 (4.10) | 24.56 (3.50) | 25.60 (4.48) |
| 2020-2023 w/o telephone interview | 83 | | 24.49 (4.13) | 25.11 (4.11) | 24.29 (4.14) |
| **Blood pressure** (high/normal) |  | |  |  |  |
| 1975 | 91 | | 6/85 | 2/36 | 4/49 |
| 1975 w/o telephone interview | 83 | | 11/72 | 4/14 | 7/58 |
| 1981 | 85 | | 13/72 | 4/34 | 9/38 |
| 1981 w/o telephone interview | 77 | | 21/56 | 6/10 | 15/46 |
| 1990 | 49 | | 11/38 | 3/14 | 8/24 |
| 1990 w/o telephone interview | 45 | | 21/24 | 4/6 | 17/18 |
| 2020-2023 | 90 | | 63/27 | 22/17 | 41/10 |
| 2020-2023 w/o telephone interview | 86 | | 54/32 | 10/11 | 44/21 |
| **Cholesterol** (high/normal) |  | |  |  |  |
| 1981 | 36 | | 7/29 | 5/18 | 2/11 |
| 1981 w/o telephone interview | 22 | | 7/15 | 3/4 | 4/11 |
| 1990 | 42 | | 23/19 | 6/10 | 17/9 |
| 1990 w/o telephone interview | 30 | | 14/16 | 2/4 | 12/12 |
| 2020-2023 | 65 | | 31/34 | 14/17 | 17/17 |
| 2020-2023 w/o telephone interview | 55 | | 17/38 | 6/11 | 11/27 |
| **Physical activity** (MET hours/day) |  | |  |  |  |
| 1975/1981 | 88 | | 2.40 (1.71) | 2.74 (1.85) | 2.14 (1.57) |
| 1975/1981 w/o telephone interview | 86 | | 1.96 (1.54) | 1.98 (1.68) | 1.95 (1.51) |
| 1990 | 52 | | 3.24 (4.56) | 4.88 (7.15) | 2.36 (1.87) |
| 1990 w/o telephone interview | 47 | | 2.41(1.98) | 2.81 (2.66) | 2.30 (1.79) |
| 2020-2023 | 84 | | 1.70 (1.65) | 1.79 (1.48) | 1.64 (1.79) |
| 2020-2023 w/o telephone interview | 54 | | 1.61 (1.80) | 2.55 (2.24) | 1.28 (1.52) |
| **CAIDE total** | 54 | | 7.61 (1.92) | 7.57 (1.73) | 7.65 (2.13) |
| **CAIDE total** w/o telephone interview | 44 | | 8.89 (2.22) | 9.7 (2.26) | 8.65 (2.19) |
| **EDU-OCU** | 94 | | 16.77 (3.15) | 16.68 (3.28) | 16.83 (3.08) |
| **EDU-OCU** w/o telephone interview | 88 | | 15.08 (2.26) | 14.86 (2.24) | 15.15 (2.28) |
| **Education 1975** (<6yrs/ 7-11 yrs./ >12 yrs.) | 82 | | 33/40/19 | 11/19/8 | 22/21/11 |
| **Education 1975** w/o telephone interview | 92 | | 55/24/3 | 10/8/0 | 45/16/3 |
| **Education 1981** (<6yrs/ 7-11 yrs./ >12 yrs.) | 94 | | 33/41/20 | 13/18/9 | 20/23/11 |
| **Education 1981** w/o telephone interview | 89 | | 58/27/4 | 12/8/0 | 46/19/4 |
| **Education 90 yrs.** (<6yrs/ 7-11 yrs./ >12 yrs.) | | 96 | 38/38/20 | 17/15/9 | 21/23/11 |
| **Education 90 yrs.** w/o telephone interview | 91 | | 62/25/4 | 15/6/0 | 47/19/4 |
| ***APOE* status** ( ε4 carrier/non-carrier) | 83 | | 20/65 | 5/31 | 15/34 |
| ***APOE* status** w/o telephone interview | 63 | | 17/46 | 5/9 | 12/37 |

APOE = apolipoprotein E, BMI = body mass index, CAIDE = Cardiovascular Risk Factors, Aging and Dementia score, EDU-OCU = educational-occupational score, MET = metabolic equivalent hours per day, SD = standard deviation, w/o = without, yrs.= years.

**S15 Table. Two-sample t-test results, examining midlife and old age cardiovascular risk factors and risk scores in those who participated in telephone interview and questionnaire at 90 years old, those who only participated in questionnaire at age 90, and those who were invited but did not participate.**

|  | **N** | | | **Telephone interview & questionnaire vs. neither** | | | | **Telephone interview & questionnaire vs. questionnaire only** | | | | **Mean (SD)** | | |
| --- | --- | --- | --- | --- | --- | --- | --- | --- | --- | --- | --- | --- | --- | --- |
| **Risk factor** | tele & qtn | qtn only | neither | t | *p* | 95% CI low | 95% CI upper | t | *p* | 95% CI low | 95% CI upper | tele & qtn | qtn only | neither |
| 75/81 BMI | 94 | 90 | 500 | 0.47 | 0.639 | -0.52 | 0.85 | -1.06 | 0.289 | -1.31 | 0.39 | 24.67 (3.05) | 24.22 (2.65) | 24.84 (2.74) |
| 1990 BMI | 51 | 48 | 198 | 0.91 | 0.366 | -0.95 | 2.56 | -0.75 | 0.456 | -1.82 | 0.83 | 25.78 (3.72) | 25.28 (2.96) | 26.59 (10.34) |
| 90 v. BMI | 95 | 83 | - | - | - | - | - | -1.06 | 0.291 | -1.89 | 0.57 | 25.15 (4.10) | 24.49 (4.13) | - |
| 75/81 PA | 88 | 86 | 469 | -0.28 | 0.777 | -0.48 | 0.36 | -1.75 | 0.082 | -0.94 | 0.06 | 2.40 (1.71) | 1.96 (1.54) | 2.34 (2.30) |
| 1990 PA | 52 | 47 | 193 | -0.69 | 0.493 | -1.88 | 0.91 | -1.15 | 0.254 | -2.26 | 0.61 | 3.24 (4.56) | 2.41 (1.98) | 2.75 (3.27) |
| 90 v. PA | 84 | 54 | - | - | - | - | - | -0.29 | 0.769 | -0.72 | 0.53 | 1.70 (1.65) | 1.61 (1.80) | - |
| EDU-OCU | 94 | 88 | 487 | -4.84 | <0.001 | -2.61 | -1.10 | -3.75 | <0.001 | -2.58 | -0.80 | 16.77 (3.15) | 15.08 (2.26) | 14.91 (2.63) |
| CAIDE | 54 | 44 | 210 | 2.50 | 0.013 | 0.17 | 1.40 | 2.82 | 0.006 | 0.38 | 2.17 | 7.61 (1.92) | 8.89 (2.22) | 8.40 (1.93) |
| CAIDE w/o EDU | 54 | 44 | 210 | 0.54 | 0.592 | -0.30 | 0.53 | 2.22 | 0.029 | 0.08 | 1.41 | 5.96 (1.36) | 6.70 (1.84) | 6.08 (1.57) |

BMI = body mass index, CAIDE = Cardiovascular Risk Factors, Aging and Dementia score, CI = confidence interval, EDU-OCU = educational-occupational score, edu = education, MET = metabolic equivalent hours per day, qtn = questionnaire, SD = standard deviation, tele = telephone interview, w/o = without. yrs. = years. Analyses adjusted for non-independence of twin data.

**S16 Table. Desing based F-test results for midlife and old age risk factors in those who participated in telephone interview and questionnaire at 90 years old, those who only participated in questionnaire at age 90, and those who were invited but did not participate.**

|  | **N** | | | **Telephone interview & questionnaire vs. questionnaire only** | | | **Telephone interview & questionnaire vs. neither** | | |
| --- | --- | --- | --- | --- | --- | --- | --- | --- | --- |
| **Risk factor** | tele & qtn | qtn only | neither | Design-based F | df1, df2 | p | Design-based F | df1, df2 | p |
| BP 1975 | 91 | 83 | 458 | 2.18 | 1, 153 | 0.142 | 1.69 | 1, 463 | 0.195 |
| BP 1981 | 85 | 77 | 400 | 3.09 | 1, 142 | 0.081 | 3.69 | 1, 418 | 0.055 |
| BP 1990 | 49 | 45 | 181 | 6.70 | 1, 79 | 0.012 | 0.82 | 1, 190 | 0.367 |
| BP 90 yrs. | 90 | 86 | - | 0.99 | 1, 155 | 0.321 | - | - | - |
| Chol 1981 | 36 | 22 | 157 | 1.17 | 1, 54 | 0.284 | 0.06 | 1, 180 | 0.813 |
| Chol 1990 | 42 | 30 | 122 | 0.43 | 1, 63 | 0.514 | 0.64 | 1, 144 | 0.426 |
| Chol 90 yrs. | 65 | 55 | - | 3.16 | 1, 110 | 0.078 | - | - | - |
| EDU 1975 | 92 | 82 | 466 | 9.55 | 2.00, 307.73 | <0.001 | 19.19 | 2.00, 941.60 | <0.001 |
| EDU 1981 | 94 | 89 | 501 | 8.05 | 1.96, 314. 95 | <0.001 | 19.59 | 2.00, 1005. 83 | <0.001 |
| EDU 90 yrs. | 96 | 91 | - | 7.82 | 1.94, 318.05 | <0.001 | - | - | - |

BP = blood pressure, Chol = cholesterol, df = degrees of freedom, EDU = education, qtn = questionnaire, tele = telephone interview. yrs. = years. Analyses adjusted for non-independence of twin data.
